# Acute infarct segmentation on diffusion-weighted image using deep learning algorithm and RAPID DWI: a comprehensive stroke center clinical validation study

**DOI:** 10.1101/2023.02.20.23286140

**Authors:** Wi-Sun Ryu, You-Ri Kang, Yoon-Gon Noh, Jong-Hyeok Park, Dongmin Kim, Byeong C. Kim, Man-Seok Park, Beom Joon Kim, Joon-Tae Kim

**Author notes:** Correspondence to: Joon-Tae Kim, MD, PhD, Department of Neurology, Chonnam National University Medical School, Chonnam National University Hospital, 42 Jebongro, Dong-gu, Gwangju, 61469, Korea. Equally contributed to the work (Wi-Sun Ryu, You-Ri Kang).

## Abstract

**Background:** Accurate infarct volume measurement requires manual segmentation in diffusion weighted image (DWI) which is time-consuming and prone to variability. We compared two DWI infarct segmentation programs based on deep learning and the apparent diffusion coefficient threshold (JBS-01K and RAPID DWI, respectively) in a comprehensive stroke center.

**Method:** We included 414 patients whose DWI were evaluated using RAPID DWI and JBS-01K. We used the Bland-Altman plot to compare estimated and manually segmented infarct volumes. We compared R-squared, root mean squared error, Akaike information criterion, and log likelihood after linear regression of manually segmented infarct volumes.

**Results:** The mean age of included patients was 70,0±12.4 years, and 60.9% were male. The median time between the last known well and a DWI was 12.4 hours. JBS-01K segmented infarct volumes were more comparable to manually segmented volumes compared to RAPID DWI. JBS-01K had a lower root mean squared error (6.9 vs. 10.8) and log likelihood (p<0.001) compared to RAPID DWI. In addition, compared to RAPID DWI, JBS-01K more correctly classified patients according to the infarct volume threshold used in endovascular treatment trials (overall accuracy 98.1% vs. 94.0%; p = 0.002). In 35 patients who received DWI prior to endovascular treatment, JBS-01K infarct volume segmentation was more closely related to manual infarct volume segmentation.

**Conclusion:** We demonstrated that a deep learning method segmented infarct on DWI more accurately than one based on the apparent diffusion coefficient threshold.

## Introduction

Diffusion weighted image (DWI) is a crucial magnetic resonance imaging (MRI) for the ischemic stroke diagnosis due to its high sensitivity to the acute lesion.^1^ Stroke infarct volume on DWI predicts patient outcome and has utility for clinical trial outcomes.^2-5^

Although CT perfusion or CT angiography-based decisions may shorten door-to-puncture time, MRI-based decisions can provide a plethora of information, such as infarct core evaluation, hemorrhagic transformation, and the existence of an old infarct.^6^ In addition, the development of a fast multimodal MRI protocol^7^ and image resolution upscaling technique^8^ may gradually permit the use of MRI-based selection for endovascular treatment.

In practical practice, DWI can be assessed rapidly and easily. Yet, due to artifacts, it should be compared with ADC.^9^ Accurate infarct volume measurement requires manual segmentation of stroke boundaries in DWI which is time-consuming and prone to variability. Currently available automatic quantification tools operate in a semi-automated manner^10, 11^ or only show a glimpse for large vessel occlusion detection in acute settings.^12^ A study comparing established software applications in terms of apparent diffusion coefficient (ADC) lesion volume revealed that volume segmentation in different software products may lead to significantly different results in the individual patient,^13^ which raises concerns regarding the use of software estimating infarct core using ADC for endovascular treatment candidates.

Accrued literature has shown promising results of deep learning algorithm in segmentation of infarct volume on DWI.^14, 15^ A recent study demonstrated that deep learning algorithm outperformed RAPID (Rapid Processing of Perfusion and Diffusion, iSchema View Inc.) in infarct segmentation on DWI.^15^ However, no study has evaluated their performance in a daily clinical practice. In the study, we compared two commercial DWI infarct segmentation programs (JBS-01K and RAPID DWI) in a comprehensive stroke center.

## Method

### Study subjects

This study was a retrospective analysis of a prospective, web-based stroke registry that was a component of the Clinical Research Collaboration for Stroke in Korea registry. From August 2020 to April 2021, we screened all ischemic stroke patients hospitalized within 7 days to a stroke center (N = 673). Among them, we excluded 249 patients who did not perform DWI (n = 5) or their infarct volume was not measured by RAPID DWI (n = 244). Further we excluded 10 patients whose DWI acquisition time occurred before their documented stroke onset.

### Ethics Statements

The institutional review boards of Chonnam National University Hospital authorized the collection of clinical information and imaging data in the Clinical Research Collaboration for Stroke in Korea registry with the objective of enhancing the quality of stroke care. The requirement for written informed consent from study participants was waived due to the anonymity of the individuals and the minimal risk to them.

### Data and Image Collection

Baseline data, including NIHSS scores, were collected from all patients, and the stroke subtypes were stratified according to the TOAST criteria after complete diagnostic profiling.^16^ Cranial DICOM images were collected and segmented by two experience vascular neurologists (JTK and WSR) and large vessel occlusion (LVO) was determined using MR angiography. We defined LVO as the occlusion of internal carotid artery or middle cerebral artery (M1 or M2). The location of an infarct was classified as supratentorial if ischemic lesions were present in the cerebral hemispheres and as infratentorial if the lesions were located in the brainstem and cerebellum. Using tertile, the infarct size was classified as small (0 - 0.87 mL), medium (0.88 - 6.13 mL), and large (6.14 - 393.4 mL).

### Infarct Segmentation Programs

Two commercially available programs were used to process the DWI. RAPID DWI is an automatic infarct segmentation program based on threshold ADC value. JBS-01K (JLK Inc.) is an automatic ischemic lesion software using 3D U-net deep learning algorithm.^15^ See the supplementary material for more details. Briefly, RAPID uses the ADC threshold (620 ×lJ10^−6^ mm^2^/s) to estimate infarcted area. The JBS-01K employs deep learning algorithm trained with approximately 8,000 DWIs in which infarct areas were manually segmented by experts.

### Statistical Analysis

Comparing baseline characteristics between included and excluded subjects using the t test, rank-sum test, or chi-squares test, as appropriate. We examined the relationship between estimated infarct volumes and manually segmented infarct volumes using Pearson ‘s correlation analysis. We used Dice similarity coefficient to evaluate the inter-rater agreement of lesion segmentation in 20 cases randomly selected. Dice similarity coefficient was calculated as follows.

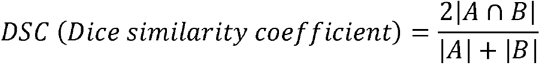

To assess the segmentation performance of JBS-01K, the dice similarity coefficient between JBS-01K and manually segmented infarct areas was determined. We used the Bland-Altman plot of automatically segmented infarct volumes by software vs manually segmented infarct volumes. In the Bland-Altman plot the percent difference was calculated as follows.

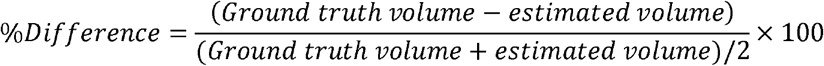

In addition, we assessed and compared R-squared, root mean squared error, Akaike information criteria, and log likelihood following linear regression analysis of automatically and manually segmented infarct volumes. We reanalyzed infarct volumes after stratification according to large vessel occlusion, infarct volume, infarct location, and time from LKW to imaging. To evaluate the accuracy of patient classification according to endovascular treatment clinical trial criteria, we categorized patients using time from LKW to image and manually segmented infarct volume and compared the frequency of correct classification between RAPID and JBS-01K using the chi-square test. In addition, for participants who received DWI prior to endovascular therapy (n = 35), we compared estimated infarct volumes by RAPID and JBS-01K to manually segmented infarct volumes using the Bland-Altman plot and parameters derived from a linear regression analysis. A two-sided p value of less than 0.05 was considered statistically significant.

## Results

### Baseline Characteristics

Among 673 patients admitted from August 2020 to April 2021, 414 patients were included for the analysis (Supplementary Figure 1). Among included patients, mean age was 70.1±12.4 years and 60.9% were men. The included patients were older, had more severe strokes, and had atrial fibrillation and cardioembolic strokes more frequently compared with excluded patients (Table 1). In addition, included patients underwent DWI earlier than excluded patients (median 12.4 hours vs. 17.7 hours; p < 0.001). Inter-rater dice similarity coefficient was 0.68 ± 0.16 and mean volume difference was 2.82 ± 4.36.

**Table 1.** Baseline characteristics comparison between included and excluded patients

|  | Included<br>(n = 414) | Excluded<br>(n = 259) | P value |
| --- | --- | --- | --- |
| Age, years | 70.1±12.4 | 68.1±12.4 | 0.04 |
| Sex, men | 252 (60.9%) | 158 (61.0%) | 0.97 |
| Admission NIHSS scores, median (IQR) | 3 (1 – 7) | 1 (0 – 4) | <0.001<sup>a</sup> |
| Previous stroke | 70 (16.9%) | 53 (20.5%) | 0.25 |
| Hypertension | 236 (57.0%) | 153 (59.1%) | 0.60 |
| Diabetes | 136 (32.9%) | 75 (29.0%) | 0.29 |
| Hyperlipidemia | 61 (14.7%) | 41 (15.8%) | 0.70 |
| Current smoking | 101 (24.4%) | 69 (26.6%) | 0.51 |
| Coronary artery disease | 52 (12.6%) | 30 (11.6%) | 0.71 |
| Atrial fibrillation | 109 (26.3%) | 45 (17.4%) | 0.007 |
| Subtype |  |  | 0.003 |
| Large artery atherosclerosis | 125 (30.2%) | 107 (41.3%) |  |
| Small vessel occlusion | 57 (13.8%) | 27 (10.4%) |  |
| Cardioembolism | 89 (21.5%) | 32 (12.4%) |  |
| Undetermined | 138 (33.3%) | 87 (33.6%) |  |
| Other-determined | 5 (1.2%) | 6 (2.3%) |  |
| Time from LKW to admission, hours, median (IQR) | 8.8 (3.0 – 20.5) | 12.6 (3.4 – 44.8) | 0.001<sup>a</sup> |
| Time from LKW to image, hours, median (IQR) | 12.4 (5.2 – 28.3) | 17.7 (7.9 – 56.4)<sup>b</sup> | <0.001<sup>a</sup> |
| Intravenous thrombolysis | 52 (12.6%) | 22 (8.5%) | 0.10 |
| Endovascular treatment | 40 (9.7%) | 20 (7.7%) | 0.39 |

Data were presented as mean±SD, median (interquartile range), or number (percentage).
<sup>a</sup>Rank-sum test was used.
<sup>b</sup>Data were missing in 15 patients.
NIHSS=National Institute Health Stroke Scale; IQR=interquartile range; LKW=last known well

### Infarct segmentation performance by deep learning algorithm versus RAPID

Both RAPID DWI and JBS-01K calculating infarct volume were significantly associated with manual segmentation infarct volume using Pearson ‘s correlation analysis (both rho = 0.98). Dice similarity coefficient between JBS-01K and manual segmentation was 0.74. Dice similarity coefficient tended to increase as infarct volume increases (Supplementary Figure 2). The segmented infarct volume dot plot revealed that volumes segmented by JBS-01K were more closely related to volumes segmented manually compared with RAPID DWI (Figure 1). RAPID DWI tended to underestimate the true infarct volume in infarct volumes less than 10 mL when compared to JBS-01K. (the inlet in Figure 1). In addition, RAPID DWI and JBS-01K were unable to detect and segment infarct in 254 (61.6%) and 8 (1.9%; p < 0.001) of 412 patients with apparent infarct on DWI, respectively. The Bland-Altman plot also indicated that JBS-01K assessed infarct volume more accurately than RAPID DWI, regardless of infarct volume. In all patients, the mean percent difference between JBS-01K and the ground truth was 20.3% (95% confidence interval [CI], -77.4 to 118.1%), while the mean percent difference between RAPID DWI and the ground truth was 110.5% (95% CI, -113.6 to 334.6%).

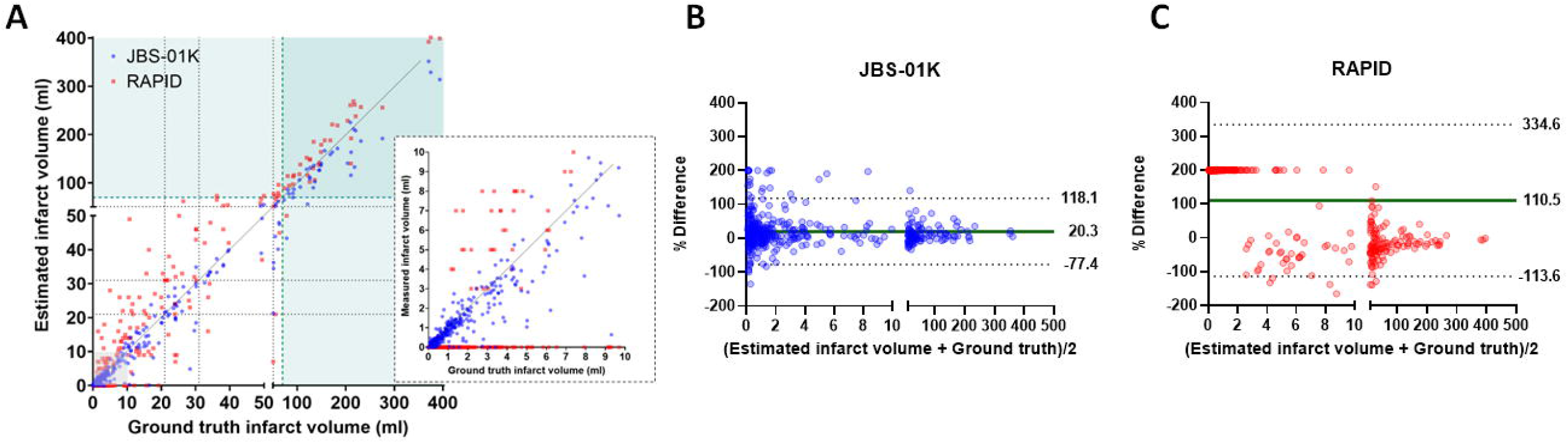

### Infarct segmentation performance by deep learning algorithm versus RAPID stratified by large vessel occlusion, infarct volume, time from last known well to imaging, and infarct location

In all patients, JBS-01K demonstrated a lower root mean squared error (Table 2; 10.76 versus 6.90) and log likelihood (p < 0.001) than RAPID DWI. After stratification by large vessel occlusion, JBS-01K predicted infarct volume more accurately than RAPID DWI in both groups with and without large artery occlusion. In the group with small infarcts (< 0.87 mL), RAPID DWI was unable of estimating infarct volume. In both the medium (0.89 – 6.13 mL) and large (6.21 – 393.4 mL) infarct groups, JSB-01K produced lower root mean squared error and log likelihood than RAPID DWI. In supratentorial or mixed and infratentorial infarcts, JSB-01K predicted infarct volume compared to manually segmented infarct volume demonstrated lower root mean square errors and log likelihood than RAPID DWI. In the < 6 hours group, infarct volume segmented by JBS-01K tended to underestimate infarct volume, whereas RAPID DWI overestimated or was unable to identify infarct (Figure 2A). In patients undergoing DWI between 6 – 24 hours from LKW, infarct volume segmented by JBS-01K differs markedly less from manual segmentation than RAPID DWI (Figure 2B). In the > 24 hours group, JBS-01K segmenting infarct volume was much more correlated to manual segmenting infarct volume than RAPID DWI.

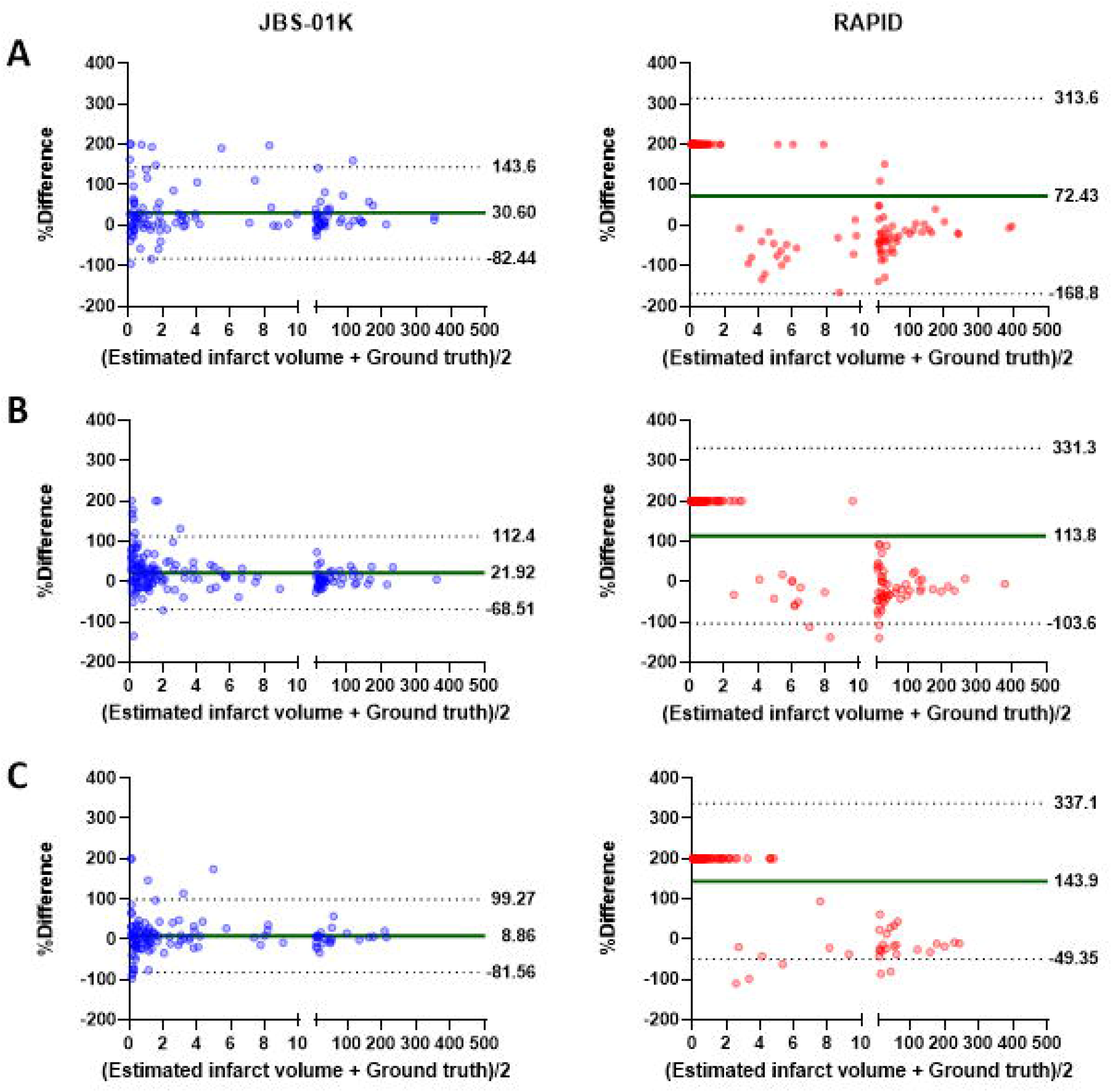

**Table 2.**
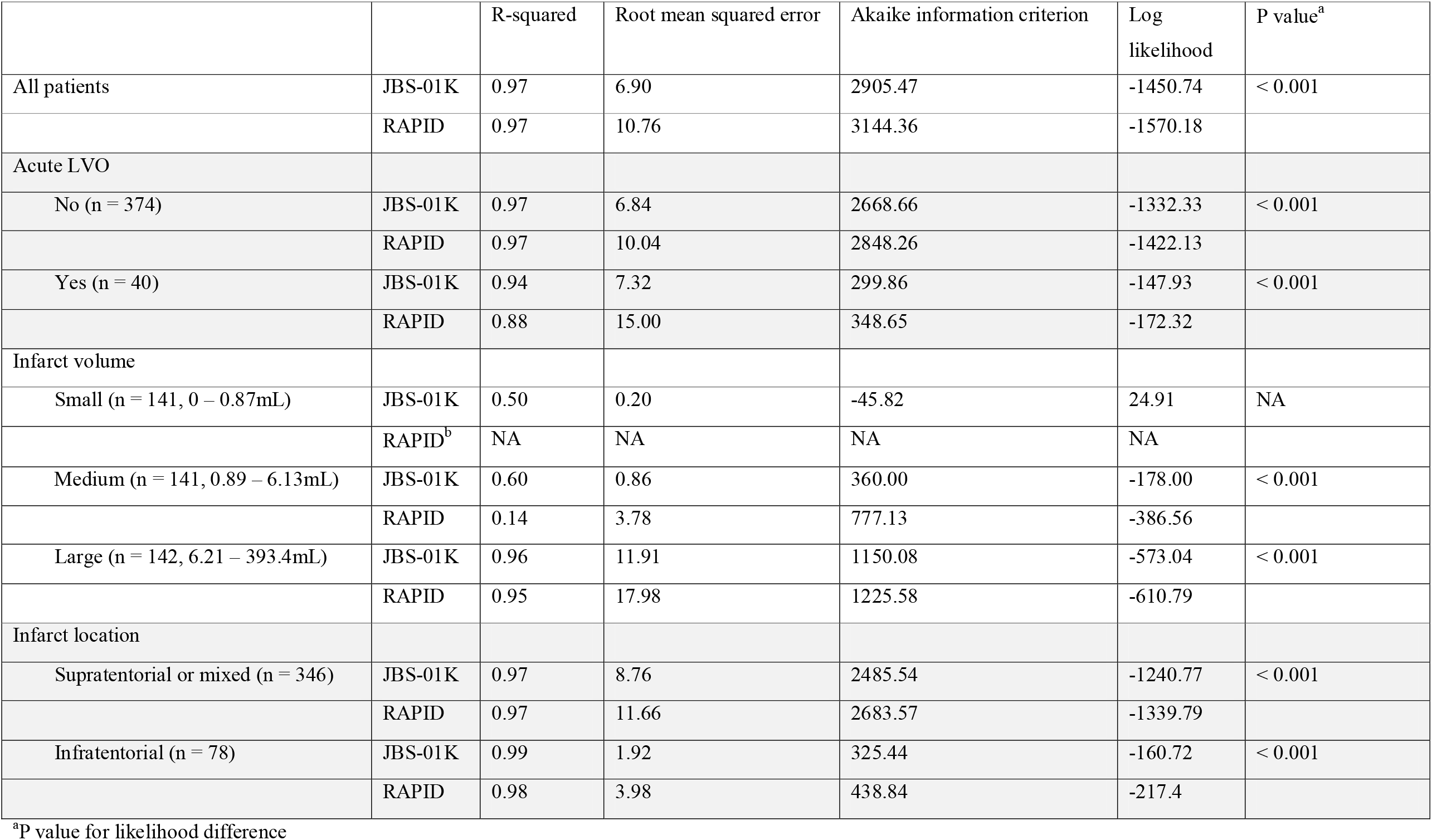
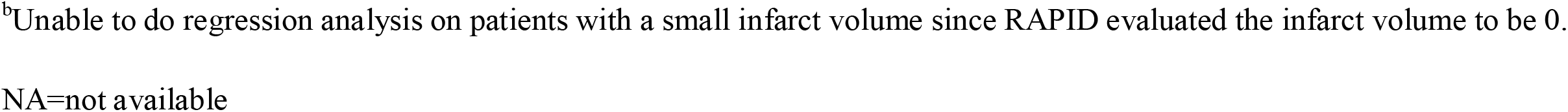
Comparison between JBS-01K and RAPID stratified by large vessel occlusion, infarct volume, and location using linear regression

|  |  | R-squared | Root mean squared error | Akaike information criterion | Log likelihood | P value<sup>a</sup> |
| --- | --- | --- | --- | --- | --- | --- |
| All patients | JBS-01K | 0.97 | 6.90 | 2905.47 | -1450.74 | < 0.001 |
|  | RAPID | 0.97 | 10.76 | 3144.36 | -1570.18 |  |
| Acute LVO |  |  |  |  |  |  |
| No (n = 374) | JBS-01K | 0.97 | 6.84 | 2668.66 | -1332.33 | < 0.001 |
|  | RAPID | 0.97 | 10.04 | 2848.26 | -1422.13 |  |
| Yes (n = 40) | JBS-01K | 0.94 | 7.32 | 299.86 | -147.93 | < 0.001 |
|  | RAPID | 0.88 | 15.00 | 348.65 | -172.32 |  |
| Infarct volume |  |  |  |  |  |  |
| Small (n = 141, 0 – 0.87mL) | JBS-01K | 0.50 | 0.20 | -45.82 | 24.91 | NA |
|  | RAPID<sup>b</sup> | NA | NA | NA | NA |  |
| Medium (n = 141, 0.89 – 6.13mL) | JBS-01K | 0.60 | 0.86 | 360.00 | -178.00 | < 0.001 |
|  | RAPID | 0.14 | 3.78 | 777.13 | -386.56 |  |
| Large (n = 142, 6.21 – 393.4mL) | JBS-01K | 0.96 | 11.91 | 1150.08 | -573.04 | < 0.001 |
|  | RAPID | 0.95 | 17.98 | 1225.58 | -610.79 |  |
| Infarct location |  |  |  |  |  |  |
| Supratentorial or mixed (n = 346) | JBS-01K | 0.97 | 8.76 | 2485.54 | -1240.77 | < 0.001 |
|  | RAPID | 0.97 | 11.66 | 2683.57 | -1339.79 |  |
| Infratentorial (n = 78) | JBS-01K | 0.99 | 1.92 | 325.44 | -160.72 | < 0.001 |
|  | RAPID | 0.98 | 3.98 | 438.84 | -217.4 |  |

<sup>a</sup>P value for likelihood difference
<sup>b</sup>Unable to do regression analysis on patients with a small infarct volume since RAPID evaluated the infarct volume to be 0.
NA=not available

### Infarct estimation with respect to endovascular treatment decision

Accuracy of patient classification in the context of DEFUSE-3 (Endovascular Therapy Following Imaging Evaluation for Ischemic Stroke), DAWN (DWI or CTP Assessment With Clinical Mismatch in the Triage of Wake-Up and Late Presenting Strokes Undergoing Neurointervention With Trevo), or EXTEND-IA (Extending the Time for Thrombolysis in Emergency Neurological Deficits With Intra-Arterial Therapy) was elaborated in supplementary table 1. Overall classification accuracy of JBS-01K was superior to RAPID DWI (p = 0.002 by chi-square). When applying DWAN threshold, the classification accuracy of JBS-01K was higher than RAPID (p = 0.007 by chi-square).

### Infarct segmentation performance by deep learning algorithm versus RAPID in patients undergoing endovascular treatment

Among 35 patients who underwent DWI before endovascular treatment, RAPID DWI was unable to predict infarct volume in 7 (20%) patients; whereas JBS-01K was unable to estimated infarct volume in 2 (5.7%). RAPID DWI tended to overestimate infarct volume whereas JBS-01K underestimated infarct volume (Figure 3). However, JBS-01K segmenting infarct volume was more closely related to manual segmenting infarct volume. Moreover, in patients with smaller infarct volume (< 10 mL by manual segmentation), JBS-01K markedly outperformed RAPID DWI (the inlet in Figure 3A). Simple liner regression against manual segmenting infarct volume also showed that JBS-01K had smaller rood mean squared error and log likelihood (p < 0.001) compared with RAPID DWI (Supplementary Table 2). The Bland-Altman plot revealed that the mean percent difference was lower in RAPID than in JBS-01K; however, the 95% CI was wider in RAPID than in JBS-01K, indicating estimation inconsistency.

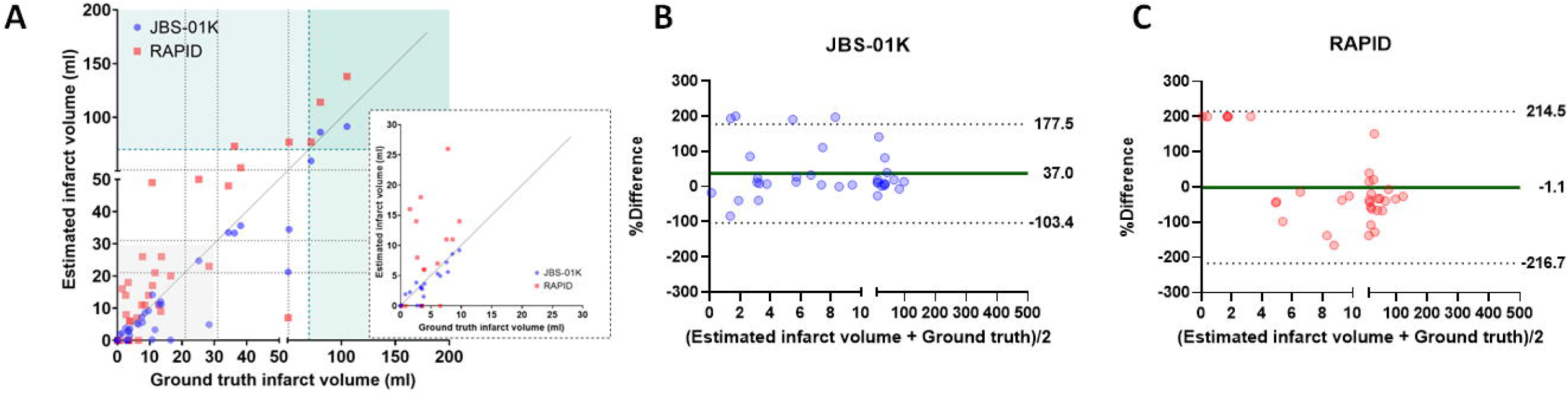

## Discussion

In acute ischemic stroke patients, we demonstrated that automated ischemic lesion segmentation on DWI by a deep learning algorithm (JBS-01K) outperformed RAPID DWI. After stratification by LKW to imaging, infarct volume, and infarct location, the deep learning method segmented infarct volume consistently better than RAPID DWI. In addition, in patients who received endovascular treatment, the deep learning algorithm segmented the infarct area more effectively than RAPID DWI. These results indicate that segmentation of lesions on DWI using a deep learning algorithm can be applied effectively in clinical practice.

In the present study, JBS-01K estimated infarct volume on DWI more accurately than RAPID DWI (Supplementary Fig 3). Notably, RAPID DWI underestimated or even failed to detect small ischemic lesion when compared to JBS-01K. In line with a recent study, it is possible that the deep learning model outperforms simple ADC thresholding by identifying ischemic lesion with subtle ADC decrease.^17^ Varying stroke pathophysiological mechanisms and stroke work flow, such as stroke subtypes, time from LKW to imaging, and age, due to different study populations across studies utilizing RAPID and ours could also account for the results.

Clinical trials on endovascular treatment for patients with large vessel occlusion have extended the intervention window for stroke,^4, 5^ increasing more patients eligible. Like a history of intravenous thrombolysis,^18^ current efforts have focused to expand the candidate pool for endovascular treatment to include distal or M2 middle cerebral artery occlusion^19, 20^ and basilar artery occlusion.^21^ As the effort continues, more precise infarct segmentation solutions that can segment both small and posterior circulation infarcts will be required. In the current study, the deep learning solution outperformed RAPID DWI in small infarct core and infratentorial stroke, indicating that, at least for MRI-guided revascularization decisions, the deep learning solution is superior to the ADC threshold-based solution.

In clinical trials and the real world, computed tomography (CT) perfusion or angiography is the mainstay due to its faster scan time.^4, 5, 22, 23^ It has been hypothesized that CT-based decision making improves clinical outcome compared to MRI-based decision making since the time between stroke onset and reperfusion is the most crucial factor in determining stroke patients ‘ outcomes.^24^ Nonetheless, accumulating evidence from real-world data has demonstrated that decisions based on MRI are not inferior to those based on CT,^22, 23, 25^ despite not having been demonstrated in randomized clinical trials. In addition, precise infarct segmentation without inter- or intra-rater variability enables investigations on serial DWIs, infarct progression prediction, and infarct–functional anatomy connection. Hence, we anticipated that a precise infarct segmentation method based on deep learning may be employed in the future to further our understanding of stroke pathophysiology utilizing DWI.

Infarct volumes on DWI correlate with functional outcome^26^ and early neurological deterioration.^27^ In addition, recent research has demonstrated that the pattern of infarct on DWI is predictive of recurrent stroke.^28, 29^ To validate and expand our understanding of DWI, a large dataset with precise infarct segmentation is required. Nevertheless, manual or semi-automated lesion segmentation is time-consuming and expensive. The technique for automated infarct segmentation demonstrated in this study may fulfill unmet needs in clinical and research fields of stroke.

Our study has a few of limitations. Patients were obtained from a single comprehensive stroke center, so it is possible that they do not accurately represent patients with suspected stroke admitted to smaller community hospitals. In the study population, a short time interval between LKW and imaging may underestimate the performance of RAPID DWI that utilized the apparent diffusion coefficient threshold. In addition, because the included patients had more severe strokes and a shorter time between LWK and imaging acquisition, the results should be interpreted with caution in the broader population of stroke patients.

In a real-world stroke cohort from a single comprehensive center, we proved that a deep learning method segmented infarct on DWI more accurately than one based on the apparent diffusion coefficient threshold. In the rapidly evolving clinical and research fields of stroke, infarct segmentation using deep learning could be a useful tool for rescuing more patients with recanalization therapy and advancing our understanding of stroke imaging.

## Data Availability

All data produced in the present study are available upon reasonable request to the authors.

## Acknowledgement

This research was supported by a grant of the Korea Health Technology R&D Project through the Korea Health Industry Development Institute (KHIDI), funded by the Ministry of Health & Welfare, Republic of Korea (grant number: HR20C0021) and the Multiministry Grant for Medical Device Development (KMDF_PR_20200901_0098) of National Research Foundation, funded by the Korean government.

