## Supplementary Material for "Acute infarct segmentation on diffusion-weighted image using deep learning algorithm and RAPID DWI: a comprehensive stroke center clinical validation study"

**Supplementary Materials**

**Supplementary methods**

**Training** data 8,661 diffusion weighted images of ischemic stroke patients from 10 university hospitals in Republic of Korea were collected. Using validated Image_QNA program, ischemic lesions were outlined by experienced researchers under and were supervised by an experience vascular neurologist.

**AI algorithm** 3D U-Net was used to train the segmentation model. To compare model performance based on training dataset size, the train and validation datasets were subsampled by a factor of 10/20/50/100%. To train the model, each subsampled dataset was divided into a train set and a validation set in 8:2 ratios. Data augmentation was used during the training process to prevent overfitting and alleviate the domain shift problem. The augmentation algorithm was written using TorchIO, a medical imaging library written in Python.

**Data augmentation.** Random augmentation was used in real time during the training process to prevent overfitting. There were four types of augmentation used: slice-wise affine transformation, MRI bias field artifact simulation, axis flip, and gamma/contrast change.

**3D U-Net.** The model consists of an encoder/decoder layer that performs max pooling four times, with the feature size set to 12, 24, 48, 96, 192 in each step (Supplementary Figure 1). The convolutional layer's kernel size is set to 3 x 3 x 3. Conv3d model weights were initialized with He normal and ConvTranspose3d model weights with Xavier uniform. For model training, the Adam optimizer was used, with batch size 2 and an exponential cyclic learning rate oscillating between 1e-5 and 1e-4. In addition, to address class imbalance caused by small lesion size compared to total brain volume, focal Tversky loss with parameters α =0.6, β =0.4, and γ= 4/3 was used.

**Environment** Python 3.7.9/3.8.13, pytorch 1.12.0, torchvision 0.13.0, pandas 1.2.4, numpy 1.19.5/1.22.3, scipy 1.4.1/1.6.3, scikit-image 0.15.0/0.18.1, SimpleITK 2.1.1, and pydicom 2.1.2 were used for all experimental procedures, including preprocessing and model development. Intel Xeon Silver 4314 @2.40GHz, 640GB RAM, and NVIDIA Quadro RTX A6000 48GB GDDR6 were used to train the models.

**Supplementary Table 1. Accuracy of patient classification according to cut off using endovascular trials**

|  | LKW to image | No. of patients | Category | JBS-01K | | | | RAPID | | |
| --- | --- | --- | --- | --- | --- | --- | --- | --- | --- | --- |
|  |  |  |  | Correct | Incorrect | Accuracy | Correct | | Incorrect | Accuracy |
| DEFUSE-3 | 6 – 16 hours | 127 | < 70 mL | 116 | 0 | 100% | 115 | | 1 | 99.2% |
|  |  |  | ≥ 70 mL | 11 | 0 |  | 11 | | 0 |  |
| DAWN | 6 – 24 hours | 171 | < 21 mL | 132 | 1 | 96.5% | 125 | | 8 | 88.9% |
|  |  |  | 21 – 31 mL | 10 | 2 |  | 6 | | 6 |  |
|  |  |  | 31- 51 mL | 5 | 1 |  | 2 | | 4 |  |
|  |  |  | ≥ 51 mL | 18 | 2 |  | 19 | | 1 |  |
| EXTEND-IA | < 4.5 hours | 86 | < 70 mL | 77 | 0 | 98.8% | 74 | | 3 | 96.5% |
|  |  |  | ≥ 70 mL | 8 | 1 |  | 9 | | 0 |  |

DEFUSE – 3 = Endovascular Therapy Following Imaging Evaluation for Ischemic Stroke; DAWN = DWI or CTP Assessment with Clinical Mismatch in the Triage of Wake-Up and Late Presenting Strokes Undergoing Neurointervention with Trevo; EXTEND-IA = Extending the Time for Thrombolysis in Emergency Neurological Deficits — Intra-Arterial; LKW = last known well.

**Supplementary Table 2. Comparison between JBS-01K and RAPID in patient undergoing endovascular treatment using linear regression (n = 35)**

|  | R-squared | Root mean squared error | Akaike information criterion | Log likelihood | P value^a^ |
| --- | --- | --- | --- | --- | --- |
| JBS-01K | 0.91 | 6.98 | 237.27 | -116.73 | < 0.001 |
| RAPID | 0.81 | 14.73 | 289.57 | -142.79 |  |

**Supplementary Figure 1. Study flow chart**

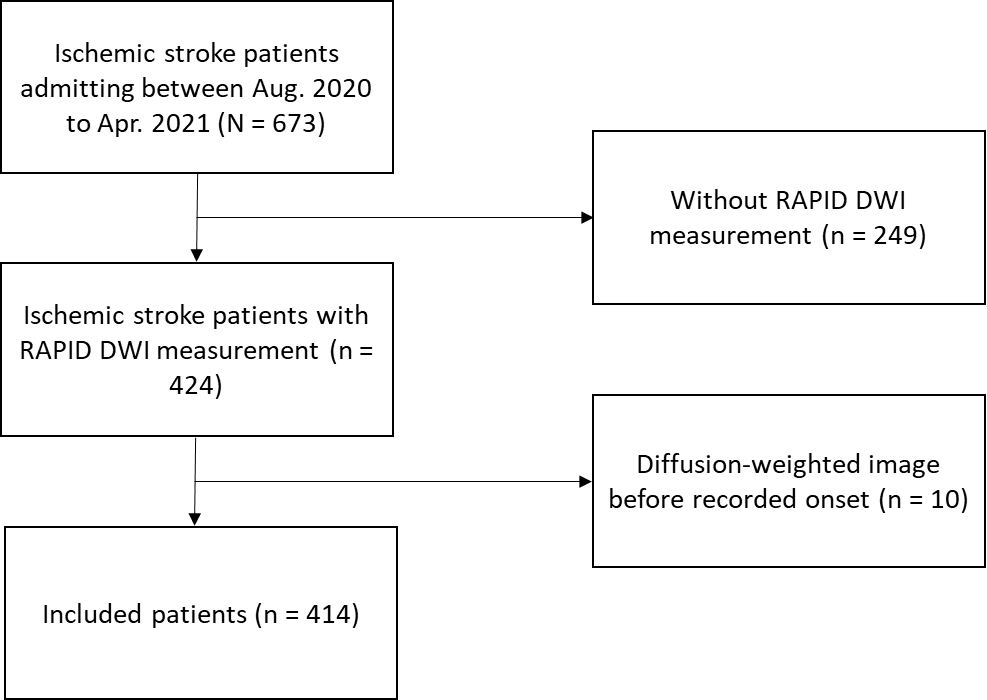

**Supplementary Figure 2. Dice similarity coefficient and ground truth infarct volume**

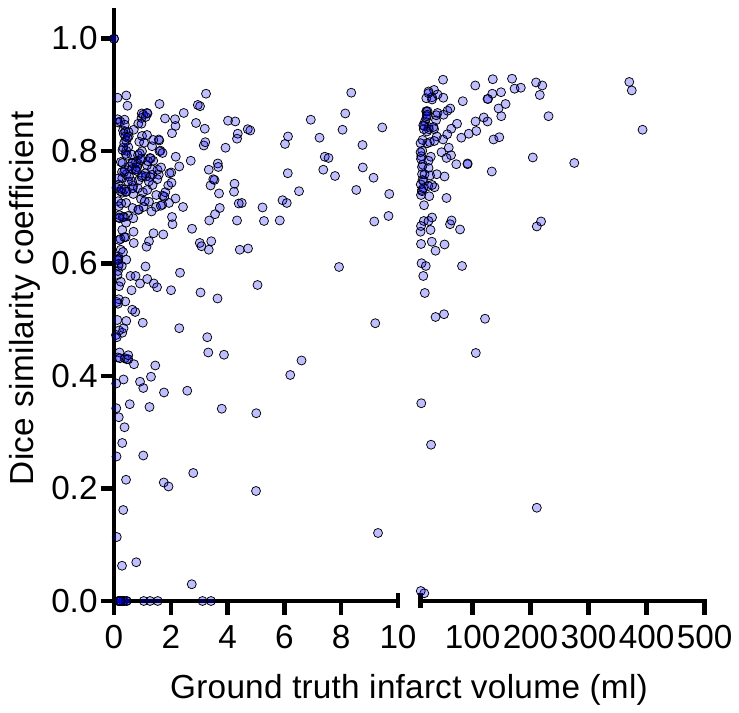

**Supplementary Figure 3. Representative cases segmented by JBS-01K and RAPID**

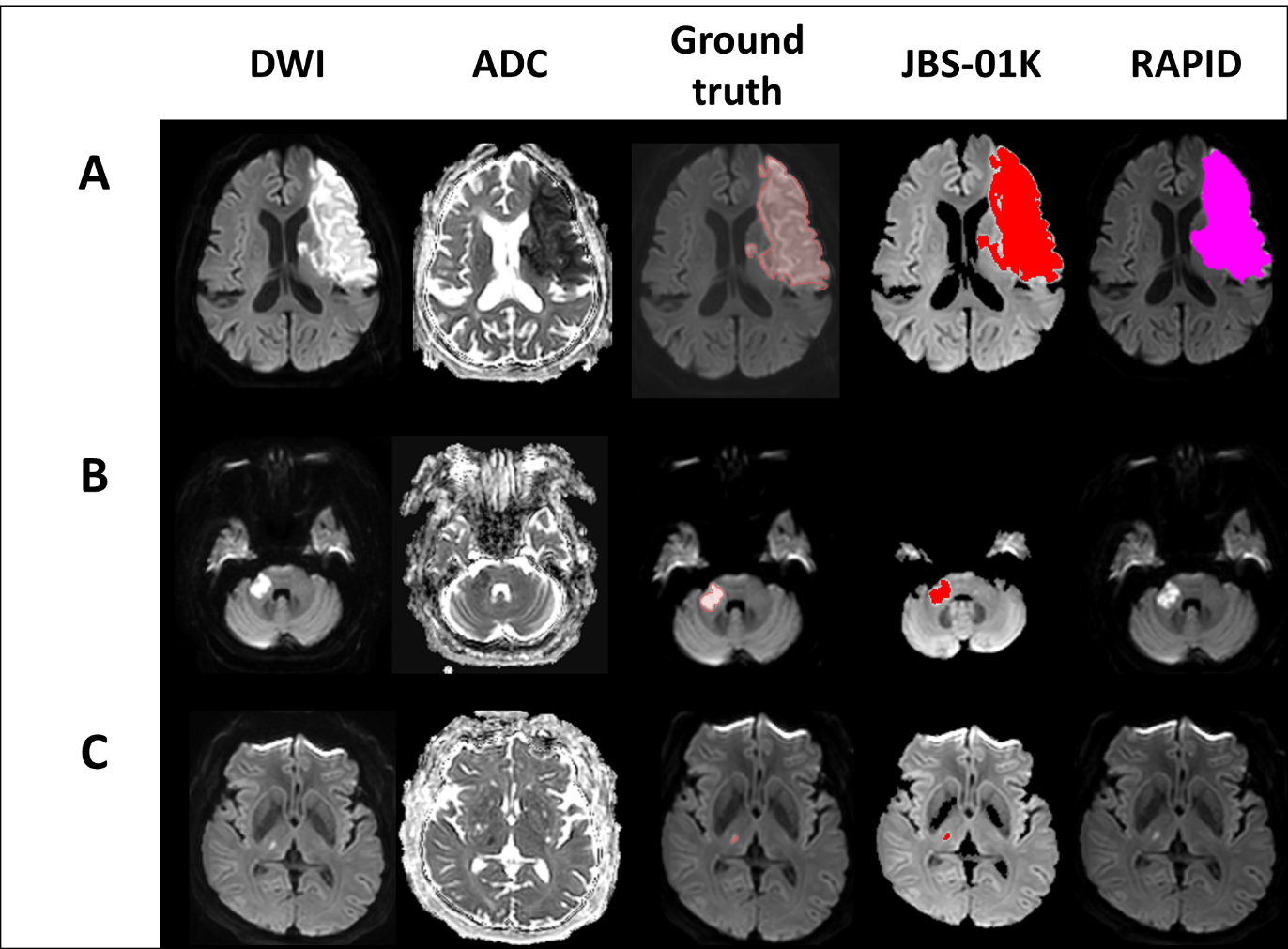

**A.** Large middle cerebral artery territory infarct. Time from last well known to image was 12.5 hours. Infarct volumes by manual segmentation, JBS-01K, and RAPID were 135, 128, and 139 mL, respectively. **B**, Infarct on middle cerebellar peduncle. Time from last well known to image was 7 hours. Infarct volumes by manual segmentation, JBS-01K, and RAPID were 2.2, 1.8 and 0 mL, respectively. **C**, Small lesion at thalamus. Time from last well known to image was 2.3 hours. Infarct volumes by manual segmentation, JBS-01K, and RAPID were 0.09, 0.07, and 0 mL, respectively.
